# Enhancing Variant of Uncertain Significance (VUS) Interpretation in Neurogenetics: Collaborative Experiences from a Tertiary Care Centre

**DOI:** 10.1101/2024.05.13.24307186

**Authors:** Kayla Horowitz, Nellie H. Fotopoulos, Alana J. Mistry, Justin Simo, Miranda Medeiros, Isabela D. Bucco, Mia Ginsberg, Emily Dwosh, Roberta La Piana, Guy A. Rouleau, Allison A. Dilliott, Sali M.K. Farhan

## Abstract

**Background:** The findings of variants of uncertain significance (VUS) on a clinical genetic testing report pose a challenge for attending healthcare professionals (HCPs) in patient care. Here, we describe the outcomes of multidisciplinary VUS Rounds, implemented at a neurological disease tertiary care centre, which aid in interpreting and communicating VUS identified in our neurogenetics patient population.

**Methods:** VUS Rounds brought together genetic counsellors, molecular geneticists, and scientists to evaluate VUS against genomic and phenotypic evidence and assign an internal temperature classification “VUS Hot”, “True VUS”, or “VUS Cold”, corresponding to potential pathogenicity. Biweekly meetings were held among the committee to deliberate variant classifications, determine additional clinical management actions, and discuss nuances of VUS result communication.

**Results:** In total, 143 VUS identified in 72 individuals with neurological disease were curated between October 2022 and December 2023. Of these, 12.6% were classified as VUS Hot, carried by 22.2% of the individuals, allowing for prioritization of additional evaluation to determine potential pathogenicity of the variants, such as clinical follow-up or segregation analysis. In contrast, 45.4% of VUS were Cold and could be eliminated from further consideration in the carrier’s care. Herein, we thoroughly evaluated the various evidence that contributed to our VUS classifications and resulting clinical actions.

**Conclusions:** The assessment of VUS leveraging multidisciplinary collaboration allowed us to delineate required follow-up analyses for our neurology patient population. Integration of VUS Rounds into healthcare practices, ensures equitable knowledge dissemination amongst HCPs and effective incorporation of uncertain genetic results into patient care.

## INTRODUCTION

In 2015, the American College of Medical Genetics and Genomics and the Association for Molecular Pathology (ACMG/AMP) established guidelines to systematically classify genetic variants as: 1) pathogenic, 2) likely pathogenic, 3) variant of uncertain significance (VUS), 4) likely benign, or 5) benign [1]. These results are reported by clinical genetic testing laboratories to the ordering healthcare professionals (HCP), who then must communicate the results to the patient. Notably, in addition to likely pathogenic and pathogenic variants, most clinical genetic testing laboratories report VUS [2 3], defined as genetic sequence changes that lack conclusive evidence to determine the biological and/or medical significance of the variant [1]. The prevalence of a VUS finding in clinical genetic test reports can be as high as 53% [4–6]. VUS necessitate HCPs provide the carrier with a nuanced explanation as to why the variant is considered uncertain and clarify implications for the individual and their family members [7]. The lack of formal genetics training among many HCPs complicates their capacity to interpret and convey these findings [8–10]. Even amongst genetic counsellors (GCs), clinical practices of returning VUS results to probands, or of recommending changes in their clinical care, vary based on individual HCP factors, such as their self-perception of their role, confidence in their skills, and practice setting [11].

VUS results demand substantial time and resources from clinicians and laboratories; pose challenges to familial testing; and complicate enrollment in clinical trials, which proves to be an emerging problem in neurogenetics precision medicine [11 12]. Due to the uncertain implications of VUS results, the recommended approach for individuals carrying a VUS is to manage their care without relying on the genetic result for clinical decision-making [1]. Falsely assigning causality to genetic findings can result in unnecessary follow-up of patients, avoidable anxiety, or false reassurance of having identified a molecular diagnosis [9 11 13]. Yet, VUS must still be effectively communicated and can be important guides for additional clinical assessment and follow-up. As such, it is repeatedly suggested that the implementation of practice guidelines, along with an incorporation of the patient’s phenotype with the VUS result and multidisciplinary collaboration between clinical specialists and genetics teams, can promote appropriate variant interpretation, precise communication of their meaning by HCPs, and suitable patient management [1 4 14].

At the academic-affiliated Montreal Neurological Institute-Hospital (The Neuro), there is an increasing incidence of non-genetics specialists receiving VUS results, which led to the development of multidisciplinary “VUS Rounds” in the Fall of 2022. An expert committee of clinical GCs, clinical molecular geneticists, and PhD- and postdoctoral-level research scientists was formed to aid in interpreting VUS identified in our neurogenetics patient population. Each variant was evaluated by the research scientists, hereafter referred to as biocurators, against genomic and phenotypic evidence types to internally assign the variant a “temperature” asserting the variant’s potential pathogenicity. Variants were categorized as either “VUS Hot,” corresponding to a variant leaning towards potential pathogenicity, “True VUS,” representing a variant that leans towards neither pathogenicity nor benignity, or “VUS Cold,” corresponding to a variant leaning towards potential benignity. Then, biweekly VUS Rounds were held among the expert committee to deliberate and confirm the final internal variant classification. The expert team also collaborated to determine potential follow-up actions in clinical management, including genetic testing of affected family members, parental segregation analysis to determine VUS phase or occurrence (inherited or *de novo*), or additional clinical investigations of the individual with neurological disease. Finally, the team discussed nuances of communicating of the VUS finding to the affected individual and to any additional HCP(s) on their clinical care team.

Here, we summarize the results of the VUS Rounds initiative from October 2022 to December 2023. This work underscores the benefits of engaging in variant curation, extending beyond the results reported by clinical genetic testing laboratories, to strengthen our understanding of the potential implications of uncertain variants. We aim to outline how multidisciplinary molecular genetics and clinical collaboration can improve the care of individuals with neurological disease and their families and enhance HCPs’ clinical workflow.

## METHODS

### Study Design

A quality improvement (QI) study design was implemented with the aim of improving VUS interpretation, enhancing patient outcomes, and promoting professional development of HCPs in cases where individuals receive uncertain neurogenetic results [15]. The project focused on refining the quality of standard patient care practices, and it did not involve procedures that posed risks to patient safety, privacy, confidentiality, or rights. Approval was obtained from The Neuro’s Director of Professional Services, and an institutional ethics review was not required.

### Identification of VUS

The study included adult individuals with neurological disease seen in Neurology at The Neuro who had undergone genetic testing between the dates of 23-Jan-2017 and 10-Nov-2023. Here, we report on VUS that were curated in VUS Rounds between October 2022 and December 2023. Providers ordering the genetic tests included neurologists and GCs, and genetic panels were ordered based on routine clinical care decisions without the consideration of VUS Rounds. All individuals provided verbal or written informed consent for genetic testing to their ordering providers.

Patient confidentiality was safeguarded by keeping personal health data, demographic data, and patient identifiers in a protected database only accessible by the certified GCs at The Neuro. Standard clinical care processes were pursued uniquely by the certified GCs, including review of patients’ medical records, communication with ordering providers and genetic testing laboratories, clinical documentation, and genetic counselling practices.

### Pre-curation of VUS

The GCs assigned all individuals with neurological disease an identification number and provided only pertinent data to the biocurators prior to the VUS Rounds, including a broad description of the patient’s phenotype; the clinical genetic testing panel ordered; the gene in which the VUS was identified and the Human Genome Variation Society (HGVS) nomenclature (coding DNA and protein change); the VUS zygosity; the gene’s associated phenotype as per Online Mendelian Inheritance in Man (OMIM) [16]; and the patient’s broad family history, if applicable.

Biocurators used information provided by the GCs to assess all lines of evidence that may influence the variant’s interpretation and classification. In addition to review of the scientific literature regarding both the VUS and the gene in which it was located, the biocurators used evidence from various resources and tools. Variant allele frequencies were obtained from general population, ancestry specific, and disease specific databases such as the Genome Aggregation Database (GnomAD) v2.1.1 and v3.1.2, including the non-neurological disease cohorts and specific ancestral subsets [17]; the Healthy Exomes dataset [18]; and the Database of Genomic Variants [19]. *In silico* prediction tools were used to assess the potential pathogenicity of variants, such as the Rare Exome Variant Ensemble Learner (REVEL) for missense variants [20] and Splice AI for splicing, intronic, or coding variants near intron-exon borders [21]. ClinVar and OMIM were used to assess previous documentations of variant pathogenicity [16 22]. While in some cases this was applicable directly to the VUS, these also identified patterns of pathogenicity within genes, such as typical sequence ontology or clustering of pathogenic/likely pathogenic variants. Finally, proteomics resources, including the InterPro [23] and the AlphaFold Protein Structure Database [24], were assessed to predict whether VUS impact protein functional domains, protein folding, or amino acid interactions. Based on all available evidence, the biocurator assigned one of three preliminary classifications: 1) “VUS Hot,” 2) “True VUS,” or 3) “VUS Cold,” as defined above.

### VUS Rounds

Following pre-curation, biweekly VUS Rounds were held among the expert committee, during which the biocurator presented all available evidence, and the GCs introduced any additional relevant clinical details. Following discussion, a final temperature classification was assigned to each VUS. The team then engaged in iterative discussions regarding whether additional clinical follow-up would clarify the role of the VUS in the carrier’s clinical picture. Follow up assessments were largely pursued for certain VUS Hot or True VUS and included magnetic resonance imaging (MRI) or blood-based analyses to identify features that may be consistent with known gene-disease relationships or segregation analysis, with the aim of gathering sufficient evidence for potential upgrade of the variant’s classification. For individuals with neurological disease carrying certain True VUS or VUS Cold, referral to ongoing research was discussed, with the hope of resolving the continued genetic diagnostic odyssey. Following Rounds, a summary of evidence and final classification for each VUS was distributed by the biocurators among the expert committee. These resources were available to share with the HCP team to enable clinical follow-up decisions and effective multidisciplinary communication. The summary resources also served as reference for HCPs when communicating the VUS results to the individuals with neurological disease.

### Data Analysis

Curated evidence for each VUS were tracked from October 2022 to December 2023. The data were manually cleaned in December 2023. For the purposes of data analysis, the clinical presentations of the individuals with neurological disease were binned across 14 phenotypic categories (**Supplemental Table 1**). In instances for which the clinical presentation of the individual may have overlapped with features across multiple bins, the phenotypic category that the individual most closely resembled was chosen. For individuals with neurological disease (n = 3) that underwent multiple clinical genetic tests, the “number of VUS identified” and “number of genes sequenced” were considered the sum of VUS identified and genes sequenced, respectively, across the genetic tests performed.

The classified VUS were retroactively analyzed to identify characteristics that influenced the final classifications, with the aim of providing insights to improve VUS interpretations in the future. The characteristics included: 1) “atypical pathogenic variant type/location,” 2) “phenotype does not match,” 3) “not enough information available,” 4) “higher than expected minor allele frequency (MAF),” 5) “incorrect inheritance pattern,” and 6) “gene-disease relationship unestablished” (definitions provided in **Supplemental Table 2**).

Finally, in January 2023, the GCs assessed the status of clinical follow-up of all individuals with neurological disease that carried VUS. Potential further clinical actions were binned into three categories: 1) segregation analysis; 2) segregation analysis and further clinical assessment; and 3) further clinical assessment. The current state of the clinical action items were defined as: 1) complete; 2) in process; 3) incomplete, referring to a conclusion following VUS Rounds by members of the clinical care team not in attendance at Rounds to not pursue the proposed clinical actions; and 4) refused, referring to a refusal by the individual with neurological disease.

Statistical analyses were performed using R statistical software v4.1.1 in R Studio v1.4.1717 [25]. Data visualization was performed using the *ggplot2* package [26]. The Sankey diagram was made using SankeyMATIC (https://sankeymatic.com/). Mean REVEL scores and GnomAD MAFs of the VUS Hot, VUS Cold, and True VUS variants were compared using pairwise Wilcoxon rank-sum tests.

## RESULTS

In total, 143 VUS identified across 72 individuals with neurological disease at The Neuro were curated between October 2022 and December 2023 (**Figure 1**). The variants spanned 110 unique genes and were commonly missense variants (n = 109, 76.2%), although frameshift insertion/deletion, non-frameshift insertion/deletion, splicing, synonymous, untranslated region, intronic, and copy number variants were also identified (**Figure 2A**).

**Figure 1.**
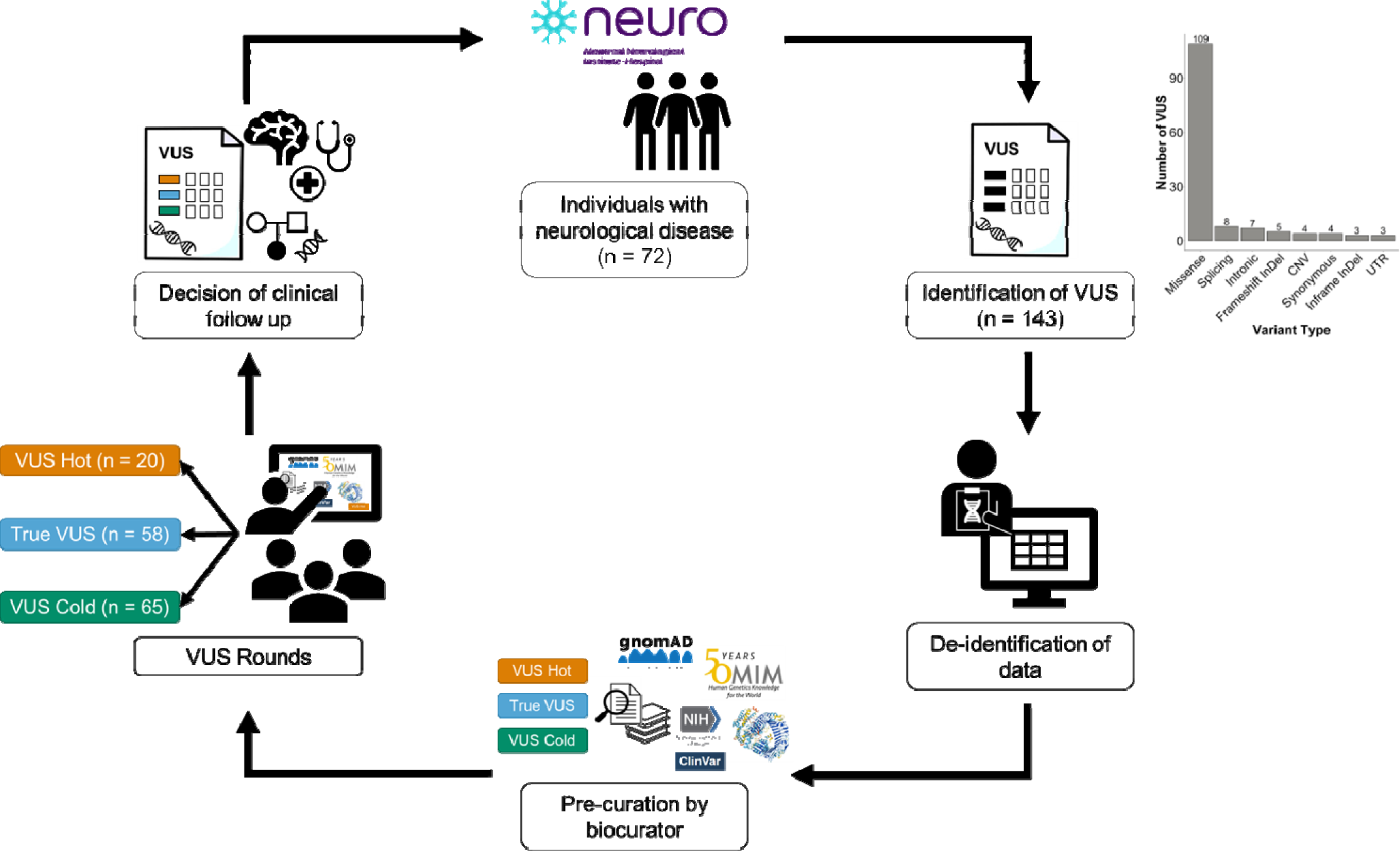
Workflow of Variant of Uncertain Significance (VUS) Rounds in a neurogenetics patient population. In total, 72 individuals with neurological disease were found to carry VUS (n = 143) through disease-specific clinical genetic testing at the Montreal Neurological Institut and Hospital. Following VUS identification, the attending genetic counsellor (GC) de-identified their phenotypic and variant details. Variant data was then pre-curated by a biocurator using a variety of resources, including population variant databases, *in silico* prediction tools, human mutation databases, genomic browsers, and scientific literature. Each VUS were assigned “temperature” and to assert the variant’s potential pathogenicity. Following preliminary variant curation, biweekly “VUS Rounds” meetings were held among the expert committee to deliberate and confirm the final internal variant classification and determine potential follow-up actions in clinical management. Following Rounds, VUS classifications were returned to the attending neurologists by the GCs and communicated appropriately to the individuals with neurological disease.

**Figure 2.**
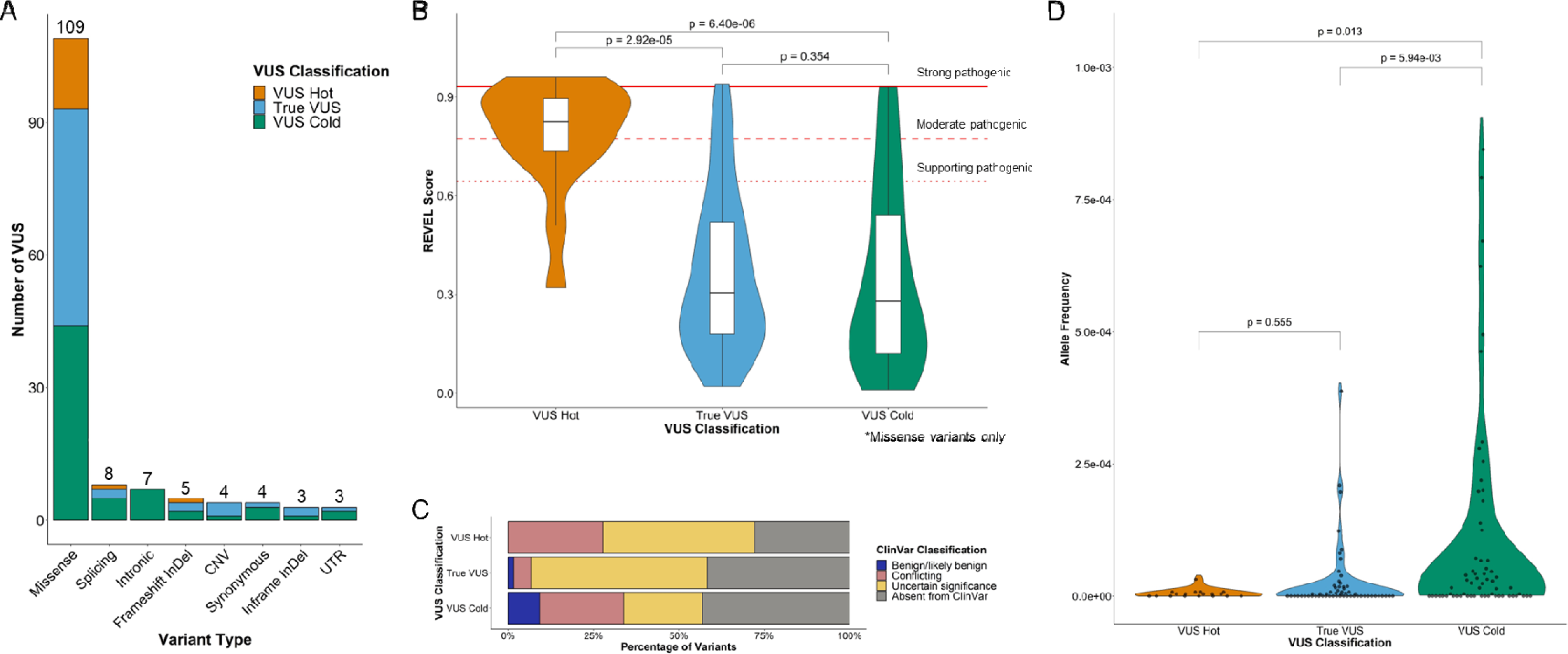
Characteristics of the 143 variants of uncertain significance (VUS) curated between October 2022 and December 2023 in VUS Rounds. The VUS were carried by 72 individuals with neurological disease from the Montreal Neurological Institute and Hospital. (A) Variant types of the 143 VUS curated in VUS Rounds. (B) Rare exome variant ensemble learner (REVEL) scores of the 109 missense variants curated in VUS Rounds. Wilcoxon’s rank sum tests were used to compare the mean REVEL scores of the variants classified as VUS Hot and VUS Cold to the variants classified as True VUS. REVEL-defined thresholds for pathogenicity are indicated by horizontal red lines. (C) ClinVar classifications of the 143 VUS curated in VUS Rounds. (D) GnomAD v2.1.1 allele frequencies of the 143 VUS curated in VUS Rounds. Variants absent from the GnomAD v2.1.1 database were assigned an allele frequency of zero for visualization purposes. Wilcoxon’s rank sum tests were used to compare the mean GnomAD v2.1.1 allele frequencies of the variants classified as VUS Hot and VUS Cold to the variants classified as True VUS.

### Factors Influencing VUS Identification and Classification

Of the 143 VUS, 18 were classified as VUS Hot (14.0%), 60 as True VUS (40.6%), and 65 as VUS Cold (45.4%). REVEL *in silico* prediction scores were used to assess the potential pathogenicity of all missense VUS. Unsurprisingly, the mean REVEL score of VUS Hot (0.785, sd = 0.170) was greater than the “moderate pathogenic” threshold defined by the method’s authors and was significantly greater than the mean REVEL score of the True VUS (0.366, sd = 0.243; p = 2.92e-05) and VUS Cold (0.342, sd = 0.278; p = 6.40e-06). Yet, the mean REVEL score of VUS Cold was not significantly different than the mean REVEL score of True VUS (p = 0.354; **Figure 2B**). When queried in ClinVar, most variants were previously reported as VUS or were absent from the database, regardless of the internal classification assigned (**Figure 2C**). No VUS had been previously reported as likely pathogenic or pathogenic in ClinVar, yet one True VUS (1.7%) and six VUS Cold (9.2%) were previously reported as likely benign or benign. Finally, the mean gnomAD v2.1.1 allele frequency of the VUS Cold (3.64e-04, sd = 1.55e-03) was significantly greater than that of the True VUS (6.65e-05, sd = 3.28e-04; p = 5.94e-03) and VUS Hot (4.65e-06, sd = 7.50e-06; p = 0.013), while the mean allele frequency of the VUS Hot was not significantly different than that of the True VUS (p = 0.555; **Figure 2D**).

Of the 72 individuals with neurological disease that carried VUS, 16 carried at least one VUS Hot (22.2%), 44 carried at least one True VUS (61.1%), and 40 carried at least one VUS Cold (55.6%; **Figure 3A**). Most VUS (n = 33) were identified in individuals with neurodegenerative disease (**Figure 3B**); although, the number of VUS identified in each phenotypic group remained generally proportional to the number of individuals in the grouping. Similarly, the VUS classifications were relatively evenly distributed across the 14 phenotypic groups.

**Figure 3.**
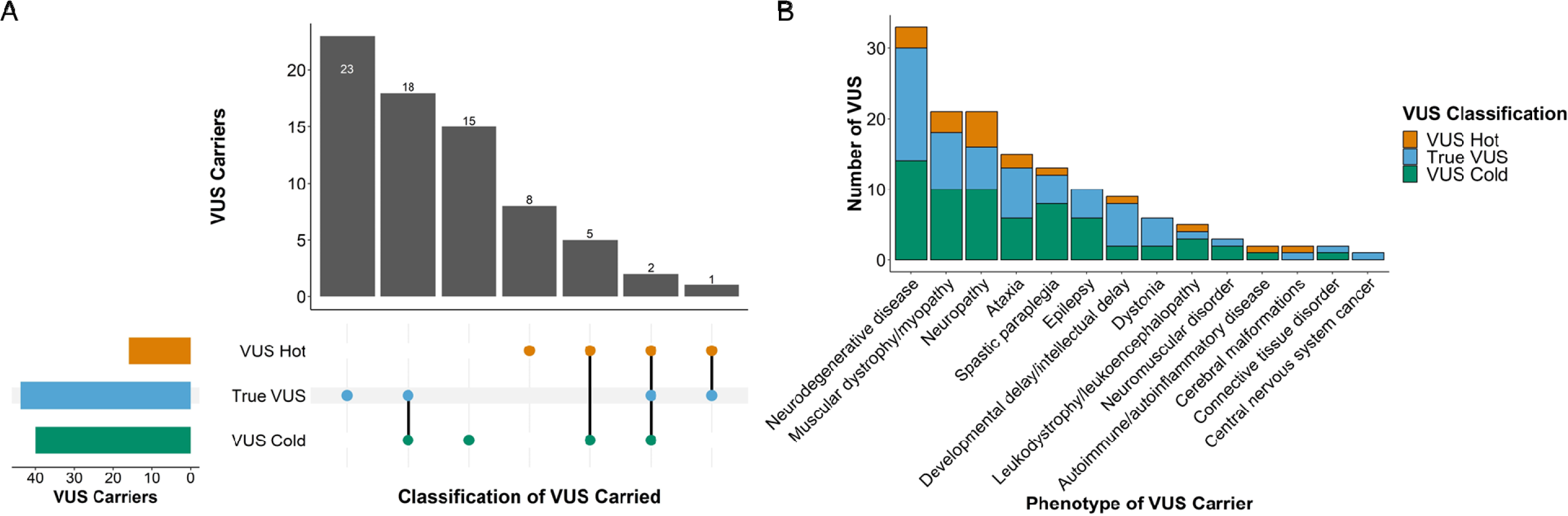
Individuals with neurological disease (n = 72) that carried the 143 variants of uncertain significance (VUS) curated between October 2022 and December 2023 in VUS Rounds. (A) VUS classifications of the variants identified through clinical genetic testing of the individual with neurological disease carriers. Notably, some patients carried multiple VUS with differing classifications, as displayed by the upset plot. The left bar plot displays the number of individuals with neurological disease carrying at least one variant of each classification type. (B) Number of VUS carried by individuals with neurological disease stratified by the general phenotype of the carrier and VUS classification.

While the phenotype of the VUS carriers was not seemingly associated with the number of identified VUS, a significant positive correlation was observed between the number of genes that sequenced and the number of VUS identified (R = 0.39, p = 0.001; **Figure 4**). One clinical genetic testing panel was excluded from this analysis, which covered >2,500 genes, as the genetic testing company indicated VUS are only reported at their discretion based on potential relevance, indicating a level of internal review.

**Figure 4.**
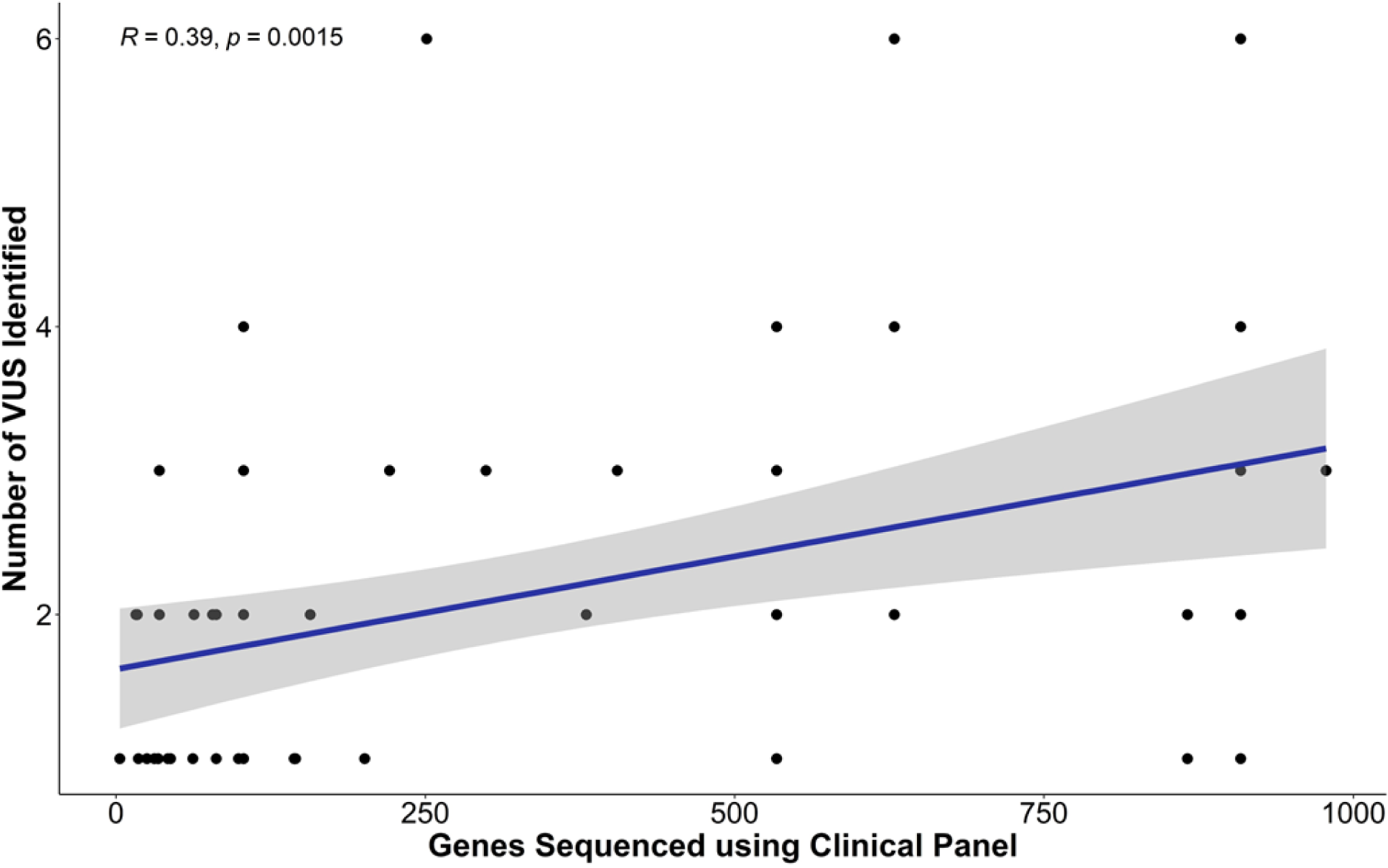
Number of variants of uncertain significance (VUS) identified per individual with neurological disease (n = 72) in relation to the number of genes that were sequenced on the clinical genetic testing panel that was ordered. The number of genes that were sequenced on the testing panel was significantly positively correlated with the number of VUS that were identified in the individual. One clinical genetic testing panel was excluded from this analysis, which covered >2,500 genes, as the genetic testing company indicated they do not return all VUS to the ordering HCPs, rather only certain VUS are returned at the company’s discretion.

We also retroactively assessed key characteristics that influenced the final VUS classifications (**Figure 5**). We found 27 of the VUS (18.9%) had insufficient evidence regarding the variant’s potential pathogenicity to drive a classification decision. Unsurprisingly, 26 were classified as True VUS. The one remaining variant was classified as a VUS Cold as it was located within a gene associated with a disease that did not align with the carrier’s presentation. Interestingly, a lack of match between the expected gene-disease association with the phenotype of the individual with neurological disease was a common characteristic, applicable to 41 VUS (28.7%). Of those, 16 were considered True VUS and 25 were VUS Cold. Other common characteristics of the classifications included the VUS being of a different sequence ontology or within a different region of the gene than previously reported pathogenic variants (n = 45, 31.5%); the VUS having an allele frequency higher than expected in general population, ancestry specific, or disease specific databases based on the disease prevalence (n = 24, 16.8%); the VUS being carried with a zygosity that does not match the expected inheritance pattern defined by the gene-disease relationship (n = 15, 10.5%); and the VUS being located within a gene without a well-established monogenic relationships with the individual’s neurological disease (n = 11, 7.69%). Largely, these characteristics were most influential in classifying variants as VUS Cold (**Figure 5**).

**Figure 5.**
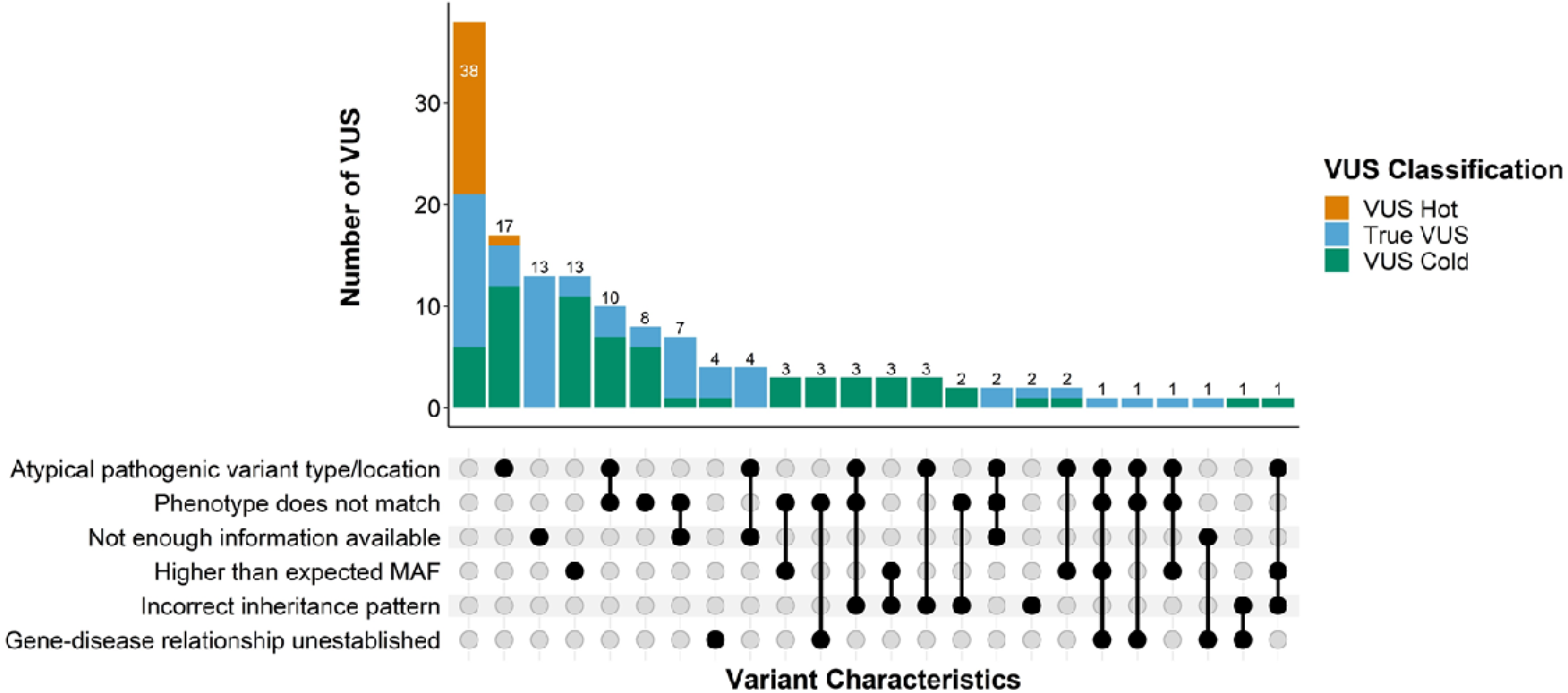
Characteristics of the 143 variants of uncertain significance (VUS) that were curated that largely drove the temperature-based classification of the variant. Although multiple lines of evidence were considered regarding each VUS, general themes from the variant curations were extracted to better understand the underlying factors impacting the VUS classification. “Atypical pathogenic variant type/location” refers to variants that were of different sequence ontology or in a different location of the gene than the majority of pathogenic variants previously reported (e.g. a missense VUS identified in a gene for which putative loss of function variants are most commonly pathogenic). “Phenotype does not match” refers to variants within genes whose established associated disease does not match the presentation of the individual with neurological disease in which the VUS was identified. “Not enough information available” refers to variants for which the variant evidence is deemed largely insufficient to drive a classification decision. “Higher than expected minor allele frequency (MAF)”’ refers to variants found at allele frequencies considered too high considering the prevalence of disease in general population, ancestry specific, or disease specific databases. “Incorrect inheritance pattern” refers to variants carried by an individual with neurological disease with a zygosity that does not match the typically expected inheritance pattern defined by the gene-disease relationship. “Gene-disease relationship unestablished” refers to variants located within genes that do not have well-established monogenic relationships with the individual with neurological disease’s presentation.

### Impact of VUS classification on patient care

Finally, we aimed to capture the impact of VUS Rounds on patient care. Of the 16 individuals with neurological disease found to carry VUS Hot, segregation analysis of the variant was pursued in unaffected family members to clarify the variant’s potential pathogenicity for 12 carriers (**Figure 6**). Segregation analysis is ongoing for six of the individuals with neurological disease, two refused the analysis, and one was not completed due to external factors. Three individuals have completed the segregation analysis, one of which also underwent additional clinical assessment relevant to the phenotype typically associated with pathogenic variants in the gene in which the VUS was located. We also pursued further clinical assessment of two VUS Hot carriers, for whom we did not choose to pursue segregation analysis (**Figure 6**). In recognition of the diagnostic odyssey still faced by the individuals with neurological disease that carried only True VUS and VUS Cold (n = 56), 15 (26.8%) were determined eligible in VUS Rounds for referral to ongoing research projects at The Neuro.

**Figure 6.**
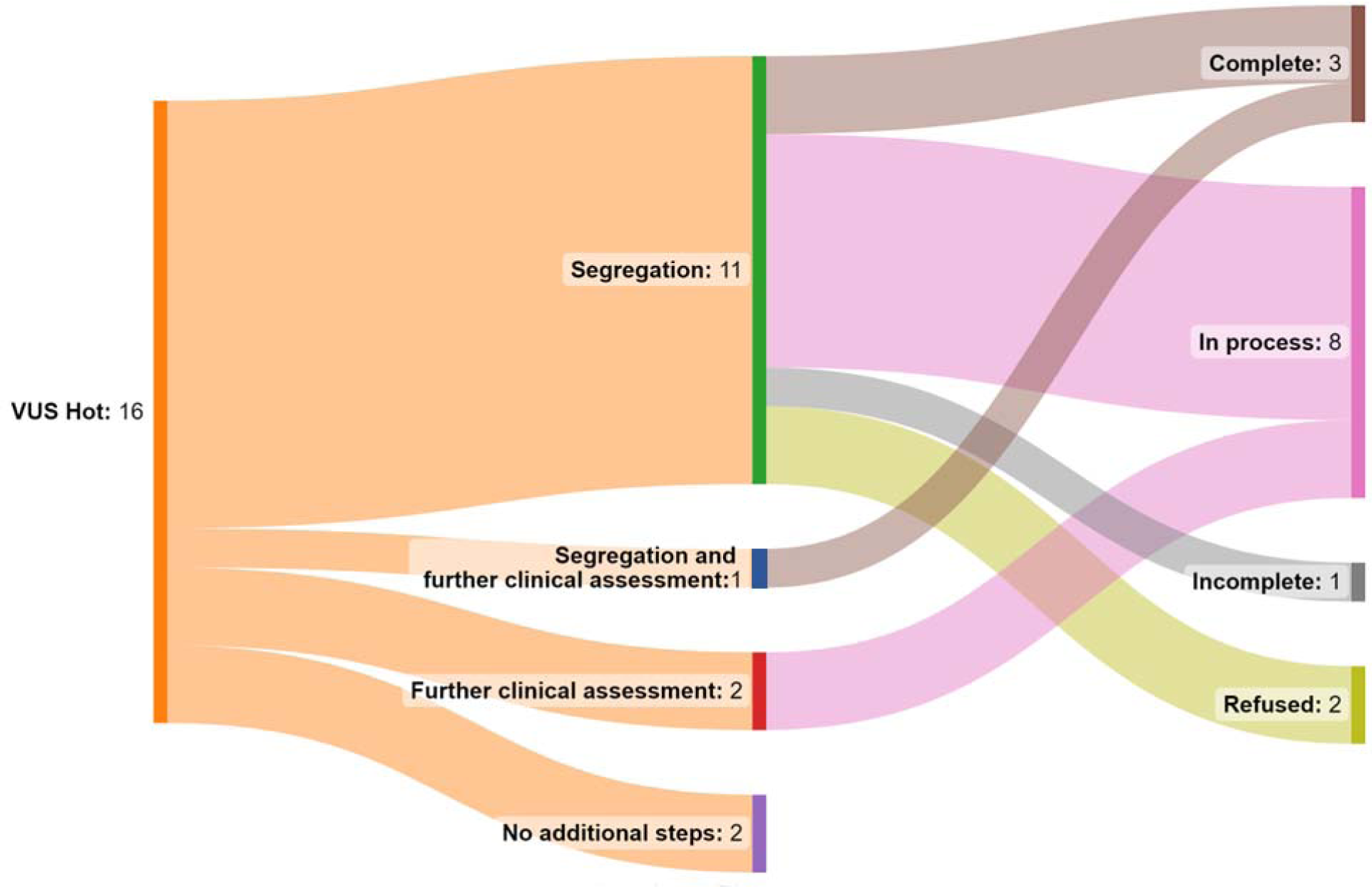
Sankey diagram depicting the decisions made within the VUS Rounds regarding clinical follow up of individuals with neurological disease carrying VUS Hot. Upon finalizing VUS classification in the Rounds setting, clinical follow up, including additional clinical assessment or segregation analysis of family members, was proposed to further confirm that the variant may be of potential interest or to improve clinical care of the carrier based on the identified VUS Hot. As of January 2023, clinical follow up was completed for three patients and was ongoing for eight patients. “Incomplete” refers to a decision following Rounds by the clinical care team to not pursue the clinical follow up proposed. “Refused” refers to a refusal for clinical follow up from the patient.

## DISCUSSION

Although the current ACMG/AMP guidelines do not recommend integration of VUS results into the clinical care of individuals with genetic disease [1], these findings are often still included on clinical genetic laboratories’ reports. In the absence of any additional pathogenic or likely pathogenic variants, communicating VUS results — and their inconclusive impact — to their carriers can be difficult for the attending clinicians and GCs. Healthcare professionals can navigate genomic uncertainties effectively by providing transparent, clear communication about the utility, actionability, and significance of genetic information [27], which have been proven to improve the psychological response of those receiving the results [5]. These considerations were driving factors for the implementation of VUS Rounds at The Neuro, as the clinical care teams sought to improve communication of uncertain results to the individuals with neurological disease and required support from those with genome analysis experience to understand the variation’s potential impact. The collaboration and clarity afforded by VUS Rounds allowed the GCs to return VUS results to the tested individuals with personalized information and assurance that a multidisciplinary team of clinical and genomic experts had comprehensively evaluated their case. Even when the results of VUS Rounds’ interpretations were communicated to the individual with neurological disease by a different HCP, the GCs could provide the outcomes from VUS Rounds to the HCP and act as a valuable source of support.

Following final multidisciplinary curation of evidence in the Rounds setting, we determined that 12.6% of the VUS were VUS Hot, carried by 22.2% of the individuals with neurological disease. Ideally, upon the identification of any VUS, further steps are taken to evaluate the potential pathogenicity of the variant, yet these may impose burden on patients and their families, and clinics may not have the resources or expertise to perform additional tests on all patients with VUS results. By curating VUS, we were able to delineate which individuals warranted additional clinical assessment or segregation analysis. Most commonly, those most appropriate for these next steps were those carrying VUS Hot. Of the 16 individuals with neurological disease carrying VUS Hot, three have had segregation testing and/or further clinical assessment completed to bolster the evidence towards the potential pathogenicity of their identified VUS. Notably, one individual carried a VUS Hot with particularly compelling combined evidence from our VUS curation, segregation testing, and additional clinical assessment. Upon recontacting the genetic testing laboratory and sharing the evidence, the laboratory agreed that the official classification of the VUS should be upgraded to likely pathogenic, resulting in an addendum to the genetic report and a genetic diagnosis for the individual with neurological disease. A previous assessment of the challenges facing genetic testing in clinics found that, generally, clinical care teams were hesitant to ask clinical testing laboratories to reclassify variants [11], yet our results highlight the benefit of pursuing these requests when the evidence supports doing so. Not only does a classification of likely pathogenic or pathogenic end the diagnostic odyssey for affected individuals, but it opens opportunities for familial predictive testing and enrollment in precision medicine-based clinical trials that are inaccessible for individuals carrying VUS [11 12].

In contrast, 45.4% of the curated VUS were VUS Cold and could be de-prioritized from further follow-up, reducing burden placed upon the individuals and their families, in addition to anticipated cost savings for the overall healthcare system. Moreover, as the majority of VUS results are eventually reclassified to benign [5], the expert committee has decided to only annually re-interpret True VUS and VUS Hot results for changes in ACMG/AMP classification, which will reduce the need for variant re-interpretation by nearly 50% in our neurological disease clinics. Finally, VUS Rounds allowed for the identification of 15 individuals with neurological disease that only carried VUS Cold or True VUS eligible to be offered enrollment in research programs, which is an avenue that we hope will further our understanding of neurogenetics and provide greater clarity for those that have not received a molecular diagnosis.

Ideally, clinical genetic testing laboratories would have access to all relevant clinical details and family history for each case, which could be essential contributors to classification workflows, yet these details are not consistently shared upon test ordering [2 14]. Unsurprisingly, these evidence types were instrumental in our curation process. For example, we found that many of the missense VUS classified as VUS Cold were either determined to be in genes for which only putative loss of function variants have been reported as pathogenic or were carried with a zygosity that did not align with the expected inheritance pattern defined by the gene-disease relationship. Our presented retrospective analysis of the characteristics that most commonly influenced VUS classification serves as a resource for future standardized guidelines for VUS interpretation and communication. Our experiences also highlight the importance of integrating the expertise of the clinical care team with molecular geneticists. Previous reports align with our findings, with one retrospective chart review of individuals that underwent clinical exome sequencing observing an increased genetic diagnostic yield of 7% following the partnership of molecular genetics laboratories with clinical teams [28].

There are many factors that influence the likelihood of identifying a VUS upon clinical genetic testing; most notably, test content. Although more comprehensive gene panels may increase pathogenic variant detection rates, they also increase the number of VUS identified [2 29 30], as corroborated by our results. We also observed that 7.69% of VUS were identified in genes that did not have well-established monogenic relationships with neurological disease. In sum, the results demonstrate the need for clinical genetic testing laboratories to be selective in the genes included in their panels. Efforts such as the NIH-funded Clinical Genome Resource (ClinGen) Gene Curation Expert Panels (GCEPs), which evaluate evidence of gene-disease relationships, will result in crucial resources moving forward to improve clinical testing panel offerings [31]. There are currently ten neurodevelopmental and neurological disorder GCEPs whose gene curations were consulted for our VUS curation efforts [12 32-34]. Recently, exome and genome sequencing were also found to decrease the rate of VUS detection in comparison to the use of multi-gene panels [2]. Largely, these results were due to the laboratories’ ability to correlate their variants with clinical and familial segregation data, again bolstering the importance of including this information, as available, when ordering genetic testing.

We recognize the presented results are specific to an institution specialized in neurological disease. Despite this, our workflow can serve as a model for implementing multidisciplinary collaboration to aid in complex genetic test result interpretation agnostic of institutional specialty. Future efforts applying this collaborative framework across diverse specialties will be imperative to ascertain if there are distinct learning needs and interpretation approaches. Moreover, our VUS curations were limited to evidence currently available through genomic resources and scientific literature. Generally, this remains a concern that must be addressed across clinical genetic efforts, as demonstrated by the well-documented increased rates of VUS in individuals from underrepresented ancestries [35]. Notably, we deemed 27 VUS as largely lacking adequate evidence to drive a classification decision, a characteristic particularly common amongst intronic, copy number, and untranslated region variants. Finally, at the conception of this QI initiative, processes to initiate variant re-interpretation and patient recontact were not defined. Despite the resource-intensive nature of these procedures, their standardization must be specified to mitigate inequities in information dissemination and health outcomes. An evaluation of the efficacy and feasibility of VUS Rounds would also be beneficial to clarify the professional hiring needs, resource allocation, and interpretation tools and databases required to adequately support variant interpretation across healthcare institutions.

## CONCLUSIONS

The use of multidisciplinary collaboration allowed us to delineate the amount and type of follow-up analysis that neurology patients may require upon receiving a VUS genetic test result. Further, our experiences highlight the opportunity for standardized VUS guidelines to be developed for clinical testing workflows. Integration of VUS Rounds into healthcare practices, particularly in non-genetics specialized centres, can ensure equitable knowledge dissemination and effective incorporation of uncertain genetic results into patient care. By systematically incorporating multidisciplinary collaboration in VUS curation, we can champion comprehensive genetic counselling and personalized clinical management, ultimately minimizing unnecessary costs to the medical system through the pursuit of suitable and effective care of each patient.

## Data Availability

The data are not available publicly due to information that could compromise the privacy of the patients. All summary data produced in the present work are contained in the manuscript

## Appendix 1: Author Contributions

| Name | Affiliations | Contribution |
| --- | --- | --- |
| Kayla Horowitz, MSc, CGC | Montreal Neurological Institute-Hospital, McGill University, Montreal, Quebec H3A 2B4, Canada | Design and conceptualization of the study; Data acquisition; Interpretation of the data; Drafting and revising of the manuscript |
| Nellie Fotopoulos, PhD, CCGC | Montreal Neurological Institute-Hospital, McGill University, Montreal, Quebec H3A 2B4, Canada | Design and conceptualization of the study; Data acquisition; Analysis of the data; Revising of the manuscript |
| Alana Mistry, MSc, CGC | Department of Human Genetics, McGill University Health Centre, Montreal, Quebec H4A 3J1, Canada | Design and conceptualization of the study; Data acquisition; Revising of the manuscript |
| Justin Simo, BSc | Montreal Neurological Institute-Hospital, McGill University, Montreal, Quebec H3A 2B4, Canada<br>Department of Neurology and Neurosurgery, McGill University, Montreal, Quebec H3A 0G4, Canada | Data acquisition; Revising of the manuscript |
| Miranda Medeiros, BSc | Montreal Neurological Institute-Hospital, McGill University, Montreal, Quebec H3A 2B4, Canada<br>Department of Human Genetics, McGill University Health Centre, Montreal, Quebec H4A 3J1, Canada | Data acquisition; Revising of the manuscript |
| Isabela Bucco, BSc | Laboratory of Human Molecular Genetics,<br>Department of Genetics,<br>Federal University of Paraná (UFPR), Centro Politécnico,<br>Jardim das Américas, Curitiba 81531-990, PR, Brazil | Data acquisition; Revising of the manuscript |
| Mia Ginsberg | Montreal Neurological Institute-Hospital, McGill University, Montreal, Quebec H3A 2B4, Canada<br>Department of Cognitive Sciences, McGill University, Montreal, Quebec H3A 2B4, Canada | Data acquisition; Revising of the manuscript |
| Emily Dwosh, CGC | University of British Columbia Hospital Clinic for Alzheimer Disease and Related Disorders, Vancouver Coastal Health, Vancouver, British Columbia, V6T 1Z3, Canada<br>Department of Medical Genetics, Faculty of Medicine, University of British Columbia, Vancouver, BC, Canada | Data acquisition; Revising of the manuscript |
| Roberta La Piana, PhD | Department of Neurology and Neurosurgery, McGill University, Montreal, Quebec H3A 0G4, Canada | Revising of the manuscript; Resources, support, and supervision |
| Guy A. Rouleau, MD, PhD | Montreal Neurological Institute-Hospital, McGill University, Montreal, Quebec H3A 2B4, Canada<br>Department of Neurology and Neurosurgery, McGill University, Montreal, Quebec H3A 0G4, Canada<br>Department of Human Genetics, McGill University, Montreal, Quebec, Canada | Revising of the manuscript; Resources, support, and supervision |
| Allison A. Dilliot, PhD | Montreal Neurological Institute-Hospital, McGill University, Montreal, Quebec H3A 2B4, Canada<br>Department of Neurology and Neurosurgery, McGill University, Montreal, Quebec H3A 0G4, Canada | Design and conceptualization of the study; Data acquisition; Analysis and interpretation of the data; Drafting and revising of the manuscript; Resources, support, and supervision |
| Sali M.K. Farhan, PhD, FCCMG | Montreal Neurological Institute-Hospital, McGill University, Montreal, Quebec H3A 2B4, Canada<br>Department of Neurology and Neurosurgery, McGill University, Montreal, Quebec H3A 0G4, Canada<br>Department of Human Genetics, McGill University, Montreal, Quebec, Canada | Design and conceptualization of the study; Data acquisition; Revising of the manuscript; Resources, support, and supervision |

## ACKNOWLEDGEMENTS

This QI initiative was undertaken in complement to standard clinical genetics workflow at The Montreal Neurological Institute and Hospital (The Neuro). No funding was acquired for the purposes of study completion. The authors thank the clinicians who refer their patients to Neurogenetics and engage in VUS Rounds for the assessment of identified VUS, including Dr. Anne-Louise Fontaine, Dr. Sandrine Larue, Dr. Rami Massie, Dr. Erin O’Ferrall, Dr. Myriam Srour, Dr. Andrea Bernasconi, Dr. Bernard Brais, Dr. Alain Dagher, Dr. Karen Dos Santos Ferreira, Dr. Edward Fon, Dr. Angela Genge, Dr. Myriam Levesque-Roy, Dr. Massimo Pandolfo, Dr. Ronald Postuma, Dr. Daniel Rabinovitch, Dr. Louise Roux, Dr. Alexander Saveriano, Dr. Madeleine Sharp, Dr. Michael Sidel, Dr. Denis Sirhan, and Dr. Chenjie Xia. R.LP. received a Research Scholar Junior 1 Award from the Fonds de Recherche du Québec – Santé (FRQS). A.A.D. is supported by the Canadian Institute of Health Research Banting Postdoctoral Fellowship program. S.M.K.F. is supported by grants from Brain Canada, ALS Society of Canada, and the Tanenbaum Open Science Institute (TOSI) at The Montreal Neurological Institute and Hospital, McGill University.

