## Supplemental Table for "Enhancing Variant of Uncertain Significance (VUS) Interpretation in Neurogenetics: Collaborative Experiences from a Tertiary Care Centre"

**Supplemental Table 1. Phenotypic bins used to categorize the diagnoses or symptoms of the individuals with neurological disease carrying VUS curated in VUS Rounds.**

| Phenotypic Bin | Individuals with neurological disease (n) |
|---|---|
| Ataxia | 7 |
| Autoimmune/autoinflammatory disease | 1 |
| Central nervous system cancer | 1 |
| Cerebral malformation | 2 |
| Connective tissue disorder | 2 |
| Developmental delay/intellectual disability | 6 |
| Dystonia | 4 |
| Epilepsy | 6 |
| Leukodystrophy/leukoencephalopathy | 3 |
| Muscular dystrophy/myopathy | 7 |
| Neurodegenerative disease | 15 |
| Neuromuscular disorder | 2 |
| Neuropathy | 9 |
| Spastic paraplegia | 7 |

**Supplemental Table 2. Definitions of the retroactively analyzed characteristics found to influence final VUS classifications.**

| Characteristic | Definition |
|---|---|
| Atypical pathogenic variant type/location | Variants of different sequence ontology or in a different location of the gene than previously reported pathogenic variants (e.g. a missense VUS in a gene for which only putative loss of function variants are pathogenic) |
| Phenotype does not match | Variants within genes whose established disease association did not match the presentation of the individual with neurological disease in whom the VUS was identified |
| Not enough information available | Variants for which the variant evidence was deemed insufficient to drive a classification decision |
| Higher than expected minor allele frequency (MAF) | Variants found at allele frequencies considered high in general population, ancestry specific, or disease specific databases based on prevalence of disease |
| Incorrect inheritance pattern | Variant with a zygosity not matching the expected inheritance pattern defined by the gene-disease relationship |
| Gene-disease relationship unestablished | Variants located within genes without well-established monogenic relationships with the individual's neurological disease (e.g. genes with only genome-wide association study (GWAS)-level evidence) |
